# Brain penetrant calcium channel blockers do not reduce alcohol consumption: Converging results from two large independent cohort studies using electronic health records

**DOI:** 10.64898/2026.02.23.26346644

**Authors:** Christopher T. Rentsch, Vanessa A. Palzes, Mingjian Shi, Michael R. Setzer, Samantha G. Malone, Andrea H. Kline-Simon, Zachary Piserchia, Emma L. Winterlind, Lorenzo Leggio, Vincent Lo Re, David A. Fiellin, John Tazare, Mehdi Farokhnia, Stacy Sterling, Henry R. Kranzler, Joshua C. Gray

## Abstract

Alcohol use disorder (AUD) remains a major public health problem, with few effective medications and suboptimal adherence. L-type calcium channel blockers (LTCCBs) have genetic and preclinical support as potential treatments for AUD. We evaluated whether brain penetrant (BP)-LTCCBs are associated with reduced alcohol consumption by conducting two preregistered (https://osf.io/huawv) observational cohort studies using electronic health records (EHRs) from the US Department of Veterans Affairs (VA) and Kaiser Permanente Northern California (KPNC). New users of BP-LTCCBs (nifedipine or felodipine) were compared with new users of a non-BP-LTCCB (amlodipine) and with unexposed patients sampled from the same clinics, following a 180-day washout and requiring at least 60 days’ supply. Propensity score matching was conducted separately for BP-LTCCB versus unexposed, non-BP-LTCCB versus unexposed, and BP-versus non-BP-LTCCB. The primary outcome was change in drinks per week from the most recent pre-index screen to end of follow-up, estimated using difference-in-differences (DiD) models. Prespecified subgroup analyses were conducted by AUD diagnosis, baseline drinking level, and sex. Across both health systems, BP-LTCCB initiation was not associated with greater reductions in drinks per week than either comparator, with broadly consistent findings across all subgroups. In two large, preregistered EHR-based cohorts with rigorous confounding control, BP-LTCCBs were not associated with reduced drinking relative to comparators. Despite compelling genetic and preclinical evidence, these results do not support repurposing BP-LTCCBs for AUD, highlighting the need to prioritize alternative pharmacologic targets, potentially within etiologically informed subgroups.

## Introduction

The 2024 National Survey on Drug Use and Health estimated that 57 million adults ages 18 and above engaged in binge drinking (i.e., ≥5 drinks on one occasion for men and 4 drinks for women) and 14.4 million reported heavy alcohol use (i.e., binge drinking on ≥5 days) in the past month^1^. Moreover, approximately 27.1 million adults were diagnosed with past-year alcohol use disorder (AUD)^1^. Individuals reporting binge drinking are more likely to have an alcohol-associated emergency department visit^2^ and 178,000 AUD-associated deaths occur each year in the US^3,4^. Despite AUD’s immense burden on public health, effective treatments are limited and underutilized.

The top medications for treating AUD include the Food and Drug Administration (FDA)-approved medications naltrexone, acamprosate and disulfiram; topiramate and gabapentin are not approved but are used off-label and endorsed by the American Psychiatric Association (APA) and other organizations^5–7^. A 2023 systematic review and meta-analysis found that oral naltrexone and acamprosate each improved alcohol consumption–related outcomes versus placebo, supporting them as first-line pharmacotherapies for AUD^8^. Meta-analyses suggest topiramate may yield larger reductions in heavy drinking than naltrexone or acamprosate, though it is off-label and often limited by adverse effects and between-trial heterogeneity^9,10^. Moreover, real-world implementation is limited: in 2024 only 2.4% of U.S. adults with past-year AUD reported received medication-assisted treatment for AUD^11^, and mean adherence across FDA-approved agents is <55% of days covered^12,13^. Considering the limited and underutilized treatment options for AUD, studies are needed to identify other potential neurobiological targets and associated treatments.

Calcium channel blockers (CCB), a class of medications used to treat hypertension (with additional use for arrhythmias, angina and coronary artery spasm), are a candidate group of widely used drugs that could be repurposed for treating AUD. Voltage-gated calcium channel subunit genes have consistently been identified in large genome-wide association studies (GWAS) of various psychiatric disorders^14–16^, and have been implicated in AUD. Recent GWAS of problematic alcohol use and alcohol consumption have identified significant variants in the voltage-gated calcium channel subunit-coding genes *CACNA1C*, *CACNA1E*, and *CACNA2D3*^17,18^. Impulsivity, smoking, and other addiction-related phenotypes have also been linked to these genes^17,19,20^.

*CACNA1C*, the calcium channel subunit gene most consistently identified in GWAS studies, encodes the L-type calcium channels, and are novel and potentially beneficial antagonist targets^17,18,21^. L-type calcium channel blockers (LTCCB) alter mesolimbic and nigrostriatal dopamine (DA) activity via plasticity-dependent changes in DA neuronal firing in the substantia nigra^22^, ventral tegmental area (VTA), and nucleus accumbens (NAc)^23^. Both VTA intracranial infusion and systemic injection of the LTCCB isradipine prevent cocaine conditioned place preference and improve conditioned place preference (CPP) extinction and reinstatement for both cocaine and alcohol^24,25^. Similarly, nimodipine, nifedipine, diltiazem, and other LTCCBs limit cocaine-, morphine-, nicotine-, and alcohol-addiction associated behaviors, such as CPP, acquisition, self-administration, progressive ratio responding, and two bottle choice in preclinical models^23,26^, an effect likely driven by the inhibition of glutamatergic synaptic plasticity in DA neurons^24^ and a reversal of VTA synaptic plasticity^27^. Research shows that LTCCBs reduce sensitization to alcohol in mice, potentially by reducing blood alcohol levels^28^.

Given evidence that medications with genetic support are more likely to be FDA-approved^29–31^ and considering support for effects of L-type CCBs on outcomes related to alcohol consumption and reward processing in preclinical studies, these medications are promising therapeutic agents that could potentially be repurposed for the treatment of AUD. LTCCBs have demonstrated safety and are FDA-approved for high blood pressure and widely used in clinical practice. A recent observational study found that a cohort of 44,731 patients prescribed brain-penetrant (BP)-LTCCBs (specifically, felodipine, isradipine, nicardipine, nifedipine, nimodipine, and nisoldipine) had a 12% reduced risk of developing mental health disorders compared to 44,731 propensity-score-matched individuals prescribed the non-BP-LTCCB amlodipine^32^. Gabapentin, which binds to the α2δ subunit of voltage-gated calcium channels, is used off-label to treat AUD, improving abstinence rates^33–35^ and reducing heavy drinking days^34,35^. Additionally, patients with a history of AUD had reduced Alcohol Use Disorders Identification Test-Consumption (AUDIT-C) scores, a quantitative measure of self-reported alcohol use, when treated with gabapentin^36^. While L-type calcium channels are genetically supported targets for psychiatric conditions^21^ with theoretical and preclinical support for treating AUD^26^, no systematic studies have examined whether BP-LTCCBs reduce alcohol consumption.

The present study used observational electronic health records (EHR) data from two large U.S. healthcare systems to evaluate associations between LTCCB receipt and changes in reported alcohol use. We hypothesized that BP-LTCCB initiators would report a greater reduction in alcohol use than non-BP-LTCCBs initiators and unexposed comparators, and this effect would be greater among individuals with AUD, those in higher AUDIT-C risk categories, and those who were prescribed higher doses. Conversely, we anticipated no sex difference in treatment effects.

## Materials and Methods

### Study Design and Data Sources

We conducted two observational cohort studies using EHR data from the US Department of Veterans Affairs (VA) and Kaiser Permanente Northern California (KPNC). We preregistered^37^ all analyses on Open Science Framework (OSF; https://osf.io/huawv) and then conducted parallel analyses in both data sources, aligning design and analytical decisions across the two studies. Both the VA and KPNC teams were blind to each other’s results until they had completed all primary analyses.

The study period was January 1, 2009, to December 31, 2023. Briefly, the VA is the largest integrated healthcare system in the US, serving ∼9 million patients annually at over 1,300 facilities nationwide. All care is recorded in the EHR with daily uploads in the VA Corporate Data Warehouse. KPNC provides care to 4.6 million members, approximately one-third of the Northern California population, at over 200 facilities. KPNC members are socio-demographically diverse and reflect the US population with access to care^38^. In KPNC, individuals enroll through various health insurance plans, including employer-based commercial plans, health insurance exchanges, Medicare, and Medicaid. Both healthcare systems offer routinely collected data on demographics, outpatient and inpatient encounters, diagnoses, laboratory results, pharmacy dispensing records, and self-reported smoking- and alcohol-related measures.

The VA study was approved by the institutional review boards of Yale University (ref #1506016006) and VA Connecticut Healthcare System (ref #AJ0013) with a waiver of informed consent. The KPNC study was approved by the KPNC Institutional Review Board (ref #1465446) with a waiver of informed consent. Both studies are compliant with the Health Insurance Portability and Accountability Act.

### Study Population and Exposure Groups

The study population comprised initiators of BP-LTCCBs and matched comparators. Three exposure groups were defined: (1) BP-LTCCBs: patients prescribed nifedipine or felodipine; (2) non-BP-LTCCBs: patients prescribed amlodipine, serving as an active comparator with limited brain penetrance; and (3) unexposed comparators: patients with no exposure to either BP- or non-BP-LTCCB. Patients prescribed co-formulations were excluded as some co-formulations may include products that impact alcohol use. We identified initiators of BP-LTCCBs or non-BP-LTCCBs, requiring a 180-day washout period to ensure new exposure episodes. Exposure to BP-LTCCBs or non-BP-LTCCBs was defined as receipt of one or more prescriptions for ≥60 days, for any indication, between January 1, 2009 and June 30, 2021, to allow all included patients two years of follow-up. The KPNC study identified exposure to BP-LTCCBs and non-BP-LTCCBs between June 1, 2013 and December 31, 2021 to align with implementation of systematic alcohol screening in primary care and allow for up to two years of follow-up. To identify the unexposed group, we identified outpatient clinics that were the largest sources of LTCCB prescriptions. We then randomly sampled from all individuals who attended at least one of these clinics–but never received a LTCCB–to ensure that unexposed patients came from the same source population, were exposed to similar medical care overall, and had an opportunity to receive an LTCCB. Index date was defined as the first dispensed date for BP-LTCCB or non-BP-LTCCB recipients and the randomly selected outpatient visit for unexposed comparators. We excluded a small number of patients who initiated BP-LTCCB and non-BP LTCCB on the same date (n=152 in VA; n=111 in KPNC).

### Inclusion and Exclusion Criteria

Patients were included if they were ≥18 years old, had at least one outpatient visit in the year prior to the index date, and had self-reported any alcohol consumption within two years prior to the index date. Due to differences in enrollment mechanisms, the KPNC study required patients to have continuous health plan membership and drug coverage (allowing up to a 60-day gap) rather than a single outpatient visit in the two years prior to the index date.

### Covariates

We extracted information on a wide range of potential confounders, harmonizing definitions and groupings across both analyses, where possible. All covariates were ascertained in the two years prior to the index date, except for co-occurring medications that were assessed as being active on the index date. We derived variables for age, sex, race, ethnicity, deprivation, urban vs rural residence, census region, and clinical characteristics including AUD, asthma, cancer, cerebrovascular disease, chronic obstructive pulmonary disease, congestive heart failure, dementia, diabetes, hemiplegia/paraplegia, human immunodeficiency virus, liver disease, mood disorder, myocardial infarction, opioid use disorder, peptic ulcer disease, peripheral vascular disease, post-traumatic stress disorder, renal disease, rheumatic disease, Charlson Comorbidity Index, and VACS Index 2.0. Deprivation was measured with the Area Deprivation Index^39^ in the VA and Neighborhood Deprivation Index^40^ in KPNC. The Charlson Comorbidity Index is a mainstay measure of overall comorbidity based on diagnostic codes across 17 clinical domains^41,42^. The VACS Index is a summary score that assesses physiologic frailty using a validated algorithm that primarily incorporates routinely available laboratory measures^43,44^. We also derived variables that captured exposure to other medications, including other CCBs, medications with demonstrated effects on alcohol consumption (such as naltrexone, acamprosate, disulfiram, gabapentin, topiramate, varenicline, spironolactone)^45–48^, any neurocognitively-active or high-burden anticholinergic medications^49^, total number of chronic medications, and site-level prescribing intensity of BP-LTCCBs. Other potential confounders included substance use treatment program visits, smoking status, body mass index, systolic and diastolic blood pressure, laboratory measures (i.e., albumin, total cholesterol, high-density lipoprotein [HDL] cholesterol, triglycerides, total bilirubin, hemoglobin, white blood cell count, fibrosis-4 [FIB-4] score, and estimated glomerular filtration rate [eGFR]); variables were categorized based on the cohort’s distribution and/or clinically relevant thresholds. We created variables denoting whether the index prescription/visit was in primary care, total number of visits to a prescribing clinic, total number of visits to any clinic, and any hospitalization in the two years prior to the index date. We created a categorical measure of BP-LTCCB prescribing level based on quartiles of the prescribing rate distribution (i.e., individuals prescribed BP-LTCCBs divided by individuals with visits), by index year and facility (VA) or service area (KPNC). Finally, baseline alcohol use was identified from the most recent alcohol screening in the two years prior to index and categorized based on the number of drinks per week using standard cut points: light (i.e., <3 for males and females), moderate (i.e., 3-14 for males or 3-7 for females), and heavy (i.e., >14 for males or >7 for females)^50^.

### Propensity Score Estimation

Propensity-score matching was performed to balance the distribution of potential confounders across exposure groups. Propensity scores (i.e., conditional predicted probabilities of exposure) were estimated using three multivariable logistic regression models, each modelling one of the three exposure contrasts of interest: BP-LTCCB vs. unexposed (A), non-BP-LTCCB vs. unexposed (B), and BP-LTCCB vs. non-BP-LTCCB (C). In the VA analysis, 56 variables described above were included in each model, of which 40 had complete data, 11 had 10-20% missingness, and all others had less than 10% missingness. Urban vs rural residence and census region were not included in the KPNC model as its population is drawn from a single region which is mostly urban. We also excluded variables from the KPNC if they had low prevalence (<1%; i.e., human immunodeficiency virus, peptic ulcer disorder, post-traumatic stress disorder), or high missingness (>20%; i.e., VACS Index, albumin, total cholesterol, HDL cholesterol, hemoglobin, total bilirubin, triglycerides, white blood cell count, FIB-4, eGFR). Of the remaining variables included in KPNC models, neighborhood deprivation index quartile, smoking status, body mass index, and systolic and diastolic blood pressure had less than 10% missingness, and all others had complete data^51,52^. We included a missing category for covariates with missing data. Under the additional assumption that associations between fully observed covariates and exposure do not differ across missingness patterns, this approach produces unbiased estimates^51,52^. Within each exposure contrast, each individual from one group was matched to one individual from the other group on the logit of the propensity score with a caliper of 0.20 times the standard deviation (SD) of the logit of the propensity score in the region of common support and using a greedy matching algorithm^53^. Individuals were exactly matched on sex, AUD diagnosis, and baseline alcohol use.

### Outcome Measures and Follow-up

Both healthcare systems conduct systematic alcohol screening in primary care. The VA has required annual AUDIT-C screening for all patients during routine health visits since 2008^54^. The AUDIT-C is a 3-item questionnaire measuring quantity/frequency of alcohol consumption and heavy episodic drinking^55^. To align with the alcohol use measure in KPNC, we converted responses from the first two items of the AUDIT-C to drinks per week using standard procedures^56^ based on drinking frequency and typical quantity. In KPNC, since June 2013, adults are asked evidence-based questions about the number of heavy drinking days in the past three months and then two questions about typical quantity and frequency of alcohol use (“On average, how many days a week do you have an alcoholic drink?” and “On a typical drinking day, how many drinks do you have?”), which are used to calculate average number of drinks per week^57^. Alcohol screening in KPNC occurs annually or more frequently (every 6 months) when alcohol use exceeds recommended limits at the prior screening^50^.

In the VA study, participants were followed for a maximum of two years from their index date or until their last VA visit, death, or June 30, 2023. In the KPNC study, participants were followed for a maximum of two years from their index date or until disenrollment from the health plan, death, or December 31, 2023. Additionally, BP-LTCCB and non-BP-LTCCB recipients were censored at medication discontinuation. To ensure equal follow-up time within matched pairs, unexposed comparators were censored at the total follow-up time of their matched exposed individual. Although evidence of any alcohol consumption at baseline was an inclusion criterion, availability of a post-index measure was not required for matching eligibility as such a restriction would not translate to an analogous prospective clinical trial.

### Statistical Analysis

Baseline characteristics were summarized using standard descriptive statistics. Absolute standardized mean differences (SMDs) were calculated to examine balance between each exposure contrast in the unmatched, matched, and final analytic cohort after restricting the matched cohorts to those with outcome measurement. We used a threshold of SMD ≤ 0.1 to indicate covariate balance, permitting values up to 0.2 for variables with small cell sizes, which can occur even when the propensity score model is correctly specified^58^. Among individuals in the final analytic cohort, we calculated average pre- and post-index drinks per week. Pre-index drinks per week was defined as the closest measure on or before the index date, within a maximum of two years prior. Post-index drinks per week was defined as the measure closest to the end of follow-up, allowing for up to 90 days after the end of follow-up. We then used multivariable difference-in-difference (DiD) linear regression models to estimate the differential change between pre- and post-index drinks per week for each exposure contrast^59,60^. Given the minimization of potential confounding by indication comparing BP-LTCCB recipients to non-BP-LTCCB recipients (contrast C), a priori, we believed the comparison would provide the most robust evidence for testing our hypothesis whether BP-LTCCBs reduce alcohol use. We also performed subgroup analyses stratified by AUD diagnosis, baseline alcohol use, and sex.

In the VA study, data management was performed using Microsoft SQL Server Management Studio v20.2, and statistical analyses were conducted using SAS Enterprise Guide v8.3 (SAS Institute, Cary, NC). In the KPNC study, all data management and statistical analyses were conducted using SAS Grid, Version 9.4 (SAS Institute, Cary, NC). All hypothesis tests were two-sided with p < 0.05 considered statistically significant.

## Results

### VA Study Population

We identified 35,825 BP-LTCCB recipients, 706,646 non-BP-LTCCB recipients, and 2,628,374 eligible unexposed comparators who reported alcohol use in the two years prior to the index date. Propensity-score matching was performed separately for each exposure contrast and resulted in 35,105 patients per group for BP-LTCCB vs. unexposed (contrast A), 504,987 for non-BP-LTCCB vs. unexposed (contrast B), and 35,821 for BP-LTCCB vs. non-BP-LTCCB (contrast C). After excluding patients with no eligible outcome measurement, the final analytic cohorts included 21,825 BP-LTCCB recipients and 19,023 unexposed patients for contrast A (**Table S1**), 313,267 non-BP-LTCCB recipients and 278,900 unexposed patients for contrast B (**Table S2**), and 18,335 BP-LTCCB recipients and 18,127 non-BP-LTCCB recipients for contrast C (**Table S3**). After propensity-score matching and restricting patients to those with a measured outcome, the distribution of baseline characteristics was balanced between groups (nearly all SMDs≤0.1 except for three variables with SMDs ≤ 0.2).

### KPNC Study Population

We identified 1,420 BP-LTCCB recipients, 43,467 non-BP-LTCCB recipients, and 2,628,374 eligible unexposed comparators who reported alcohol use in the two years prior to the index date. Propensity-score matching was performed separately for each exposure contrast and resulted in 1,418 patients per group for BP-LTCCB vs. unexposed (contrast A), 43,458 for non-BP LTCCB vs. unexposed (contrast B), and 1,354 for BP-LTCCB vs. non-BP-LTCCB (contrast C). After excluding individuals with no eligible outcome measurement, the final analytic cohorts included 600 BP-LTCCB recipients and 491 unexposed patients for contrast A (**Table S4**), 23,066 non-BP-LTCCB recipients and 21,145 unexposed patients for contrast B (**Table S5**), and 451 BP-LTCCB recipients and 387 non-BP-LTCCB recipients for contrast C (**Table S6**). After propensity-score matching and restricting patients to those with a measured outcome, the distribution of baseline characteristics was balanced between groups (most SMDs ≤ 0.1 except for 17 variables with SMDs ≤ 0.2).

### Changes in Alcohol Consumption

We found no evidence that BP-LTCCB recipients had greater decreases in drinks per week than matched non-BP-LTCCB recipients or unexposed patients in the VA study (**Table 1**; **Figure 1**) or KPNC study (**Table 2**; **Figure 1**). In the VA, pre-index and post-index drinks per week among BP-LTCCB initiators was 4.14 (standard error [SE]=0.05) and 3.14 (SE=0.05), respectively, resulting in a difference of -1.00 (SE=0.07). Pre-index and post-index drinks per week among unexposed comparators was 4.06 (SE=0.05) and 3.07 (SE=0.05), respectively, resulting in a difference of -0.99 (SE=0.07). Thus, there was no difference in changes in drinks per week between BP-LTCCB initiators and unexposed comparators (DiD: 0.00, 95% CI: -0.18, 0.19, p=0.9736). When compared head-to-head, there was no statistical evidence of a difference in a change in drinks per week between BP-LTCCB initiators and non-BP-LTCCB initiators (DiD: 0.14, 95% CI -0.06, 0.34, p=0.1644). Similar evidence was found in the KPNC study comparing BP-LTCCB initiators to unexposed comparators (DiD: 0.16, 95% CI -0.67, 0.99, p=0.7019) and to non-BP-LTCCB initiators (DiD: 0.31, 95% CI -0.48, 1.11, p=0.4395).

**Figure 1.**
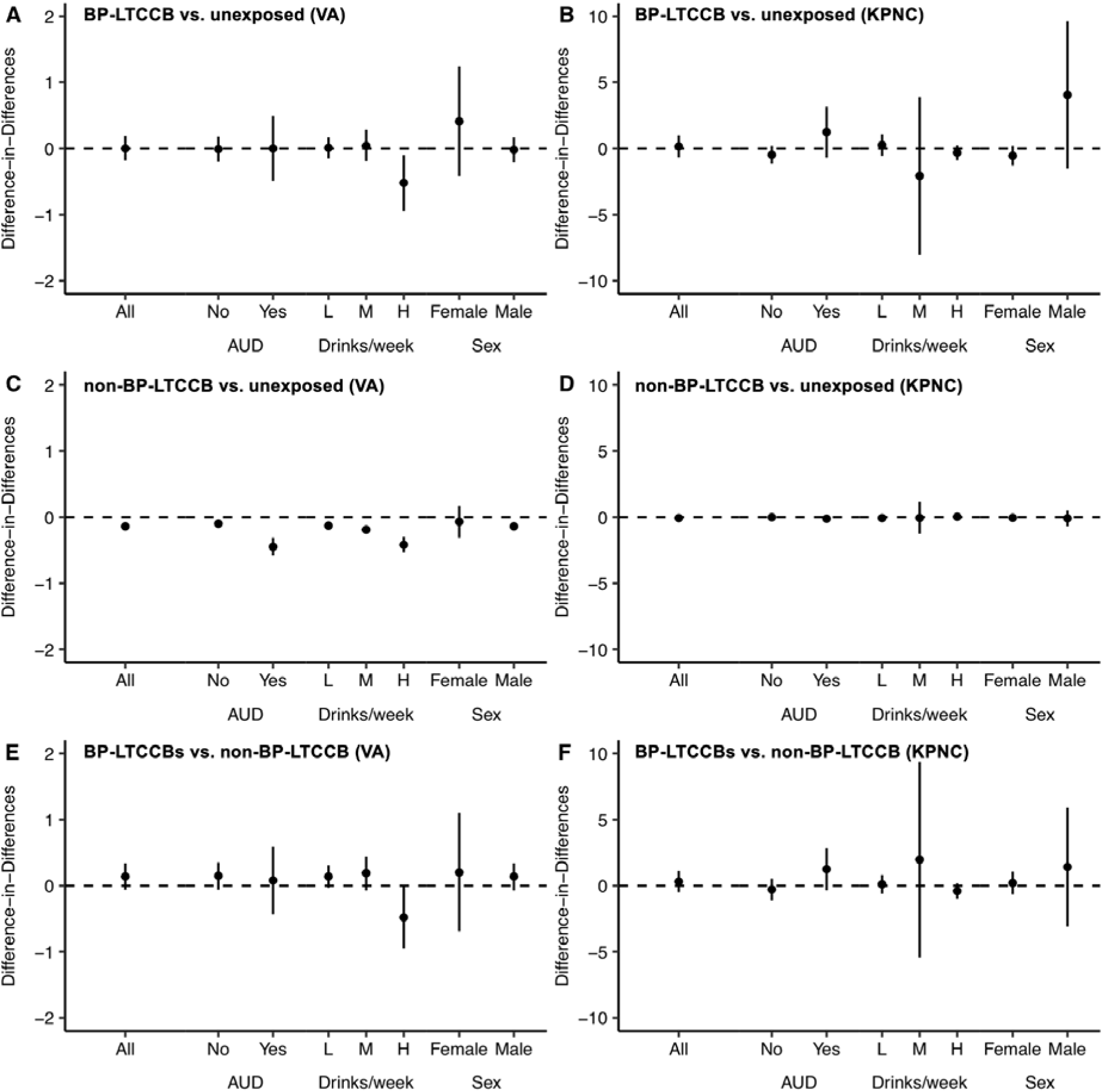
Difference-in-difference (DiD) estimates and 95% confidence intervals of self-reported changes in drinks per week overall, and stratified by baseline alcohol use disorder (AUD) diagnosis, baseline drinks per week, and sex *Abbreviations:* DiD, difference-in-difference; BP-LTCCBs, brain penetrant L-type calcium channel blocker; non-BP-LTCCBs, non-brain penetrant L-type calcium channel blocker; VA, Department of Veterans Affairs; KPNC, Kaiser Permanente Northern California; L, light (<3 drinks per week for males and females); M, moderate (3-14 drinks per week for males or 3-7 drinks per week for females; H, heavy (>14 drinks per week for males or >7 drinks per week for females).

**Table 1.**
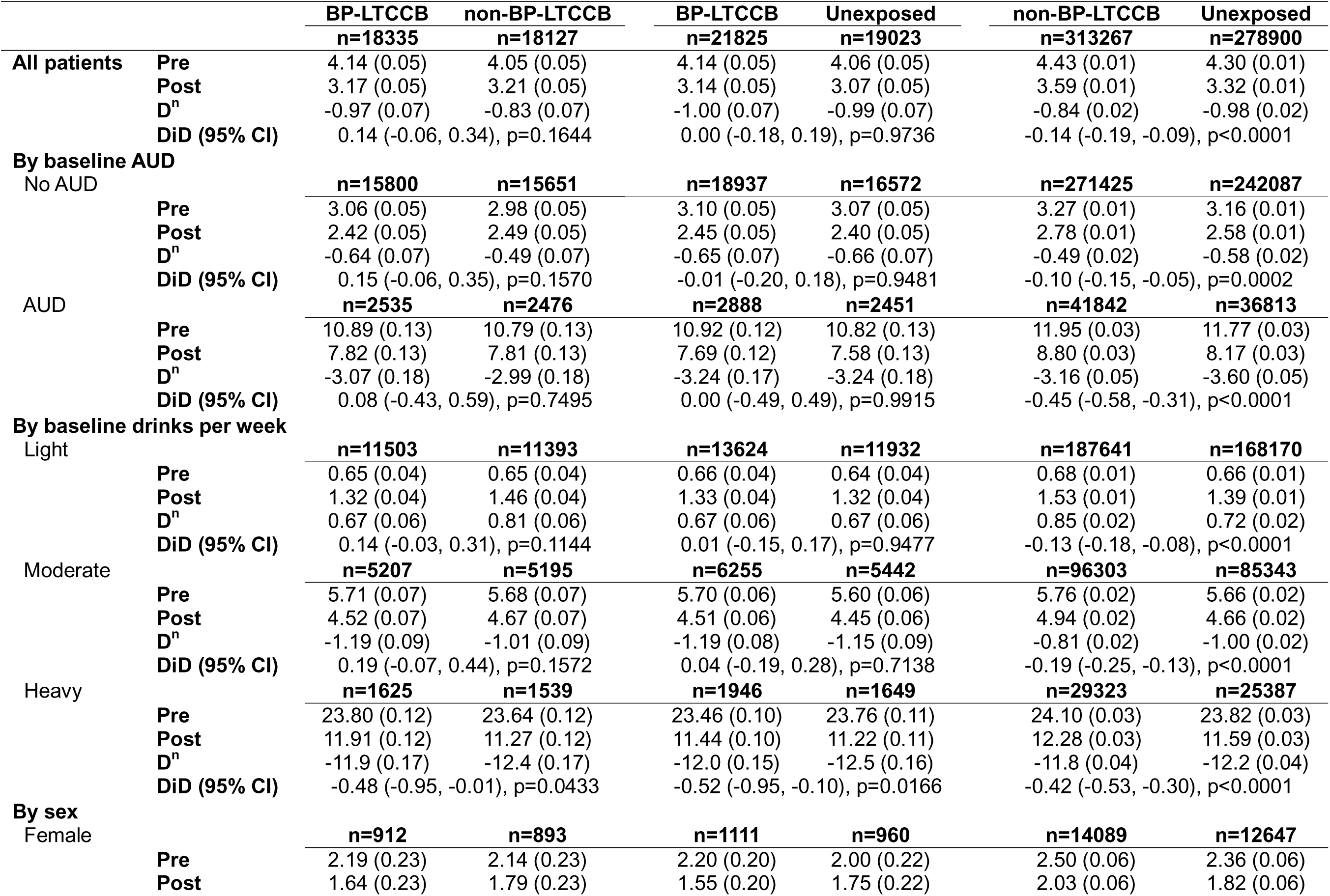
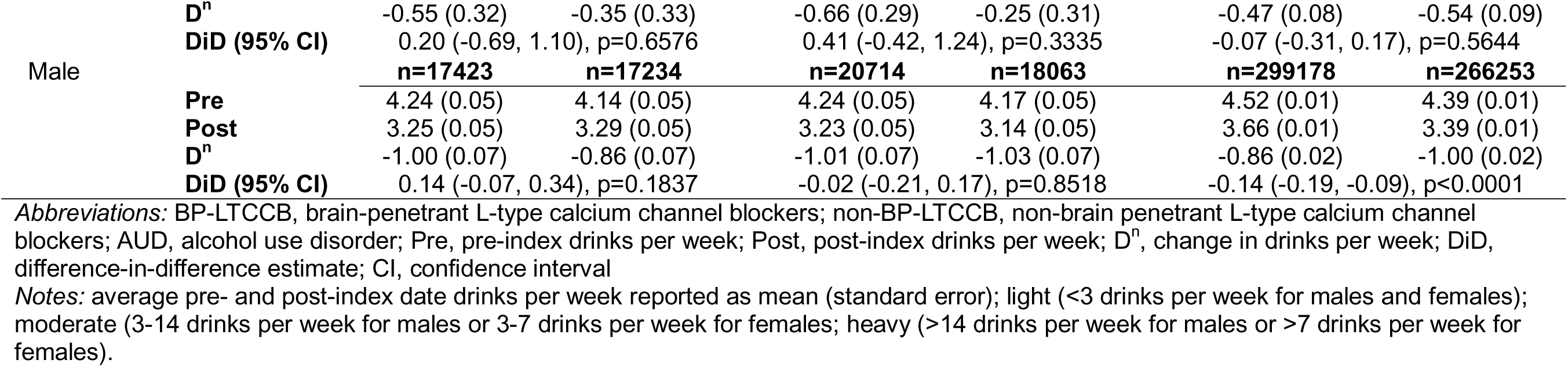
Estimated average pre- and post-index date drinks per week and difference-in-differences (DiD) across the three treatment comparisons, by baseline alcohol use disorder (AUD) diagnosis, baseline drinks per week, and sex in the VA study

**Table 2.** Estimated average pre- and post-index date drinks per week and difference-in-differences (DiD) across the three treatment comparisons, by baseline alcohol use disorder (AUD) diagnosis, baseline drinks per week, and sex in the KPNC study

|  |  | BP-LTCCB<br>n=451 | non-BP-LTCCB<br>n=387 | BP-LTCCB<br>n=600 | Unexposed<br>n=491 | non-BP-LTCCB<br>n=23066 | Unexposed<br>n=21145 |
| --- | --- | --- | --- | --- | --- | --- | --- |
| All patients | Pre | 5.64 (0.28) | 5.88 (0.30) | 5.43 (0.26) | 6.01 (0.28) | 6.99 (0.05) | 6.80 (0.05) |
|  | Post | 3.59 (0.28) | 3.51 (0.30) | 3.48 (0.26) | 3.89 (0.28) | 4.89 (0.05) | 4.74 (0.05) |
|  | D<sup>n</sup> | -2.05 (0.28) | -2.37 (0.30) | -1.96 (0.28) | -2.12 (0.31) | -2.11 (0.05) | -2.06 (0.05) |
|  | DiD (95% CI) | 0.31 (-0.48, 1.11), p=0.4395 |  | 0.16 (-0.67, 0.99), p=0.7019 |  | -0.04 (-0.17, 0.09), p=0.5213 |  |
| **By baseline AUD** |  |  |  |  |  |  |  |
| No AUD |  | n=431 | n=355 | n=574 | n=465 | n=22100 | n=20285 |
|  | Pre | 5.37 (0.25) | 5.44 (0.28) | 5.22 (0.25) | 5.83 (0.28) | 6.67 (0.04) | 6.51 (0.05) |
|  | Post | 3.37 (0.25) | 3.33 (0.28) | 3.40 (0.25) | 3.75 (0.28) | 4.76 (0.04) | 4.61 (0.05) |
|  | D<sup>n</sup> | -2.00 (0.24) | -2.11 (0.27) | -1.82 (0.28) | -2.08 (0.31) | -1.92 (0.04) | -1.90 (0.04) |
|  | DiD (95% CI) | 0.11 (-0.60, 0.82), p=0.7566 |  | 0.26 (-0.56, 1.07), p=0.5407 |  | -0.02 (-0.14, 0.10), p=0.7604 |  |
| AUD |  | n=20 | n=32 | n=26 | n=26 | n=966 | n=860 |
|  | Pre | 11.55 (2.49) | 10.72 (1.97) | 10.04 (1.67) | 9.19 (1.67) | 14.36 (0.40) | 13.75 (0.42) |
|  | Post | 8.30 (2.49) | 5.50 (1.97) | 5.12 (1.67) | 6.35 (1.67) | 7.86 (0.40) | 7.72 (0.42) |
|  | D<sup>n</sup> | -3.25 (2.89) | -5.22 (2.28) | -4.92 (2.10) | -2.85 (2.10) | -6.49 (0.46) | -6.03 (0.49) |
|  | DiD (95% CI) | 1.97 (-5.43, 9.37), p=0.5955 |  | -2.08 (-8.05, 3.90), p=0.4884 |  | -0.46 (-1.78, 0.86), p=0.4938 |  |
| **By baseline drinks per week** |  |  |  |  |  |  |  |
| Light |  | n=163 | n=130 | n=223 | n=177 | n=6235 | n=5768 |
|  | Pre | 1.47 (0.14) | 1.50 (0.16) | 1.48 (0.13) | 1.47 (0.15) | 1.47 (0.03) | 1.47 (0.03) |
|  | Post | 1.00 (0.14) | 1.43 (0.16) | 1.15 (0.13) | 1.46 (0.15) | 1.53 (0.03) | 1.53 (0.03) |
|  | D<sup>n</sup> | -0.47 (0.19) | -0.07 (0.22) | -0.32 (0.19) | -0.01 (0.21) | 0.05 (0.04) | 0.06 (0.04) |
|  | DiD (95% CI) | -0.40 (-0.97, 0.18), p=0.1743 |  | -0.32 (-0.88, 0.24), p=0.2674 |  | -0.01 (-0.11, 0.10), p=0.9151 |  |
| Moderate |  | n=237 | n=205 | n=313 | n=256 | n=13627 | n=12523 |
|  | Pre | 6.24 (0.26) | 5.98 (0.27) | 6.04 (0.22) | 5.86 (0.25) | 6.57 (0.04) | 6.47 (0.04) |
|  | Post | 4.18 (0.26) | 3.71 (0.27) | 4.06 (0.22) | 4.42 (0.25) | 5.01 (0.04) | 4.96 (0.04) |
|  | D<sup>n</sup> | -2.06 (0.30) | -2.27 (0.32) | -1.98 (0.26) | -1.44 (0.28) | -1.56 (0.05) | -1.51 (0.05) |
|  | DiD (95% CI) | 0.21 (-0.65, 1.06), p=0.6372 |  | -0.55 (-1.29, 0.20), p=0.1520 |  | -0.04 (-0.17, 0.08), p=0.4910 |  |
| Heavy |  | n=51 | n=52 | n=64 | n=58 | n=3204 | n=2854 |
|  | Pre | 16.22 (1.45) | 16.40 (1.44) | 16.25 (1.45) | 20.55 (1.52) | 19.52 (0.20) | 19.04 (0.21) |
|  | Post | 9.14 (1.45) | 7.90 (1.44) | 8.73 (1.45) | 8.98 (1.52) | 10.88 (0.20) | 10.26 (0.21) |
|  | D<sup>n</sup> | -7.08 (1.61) | -8.50 (1.60) | -7.52 (1.94) | -11.57 (2.04) | -8.64 (0.22) | -8.77 (0.24) |
|  | DiD (95% CI) | 1.42 (-3.08, 5.92), p=0.5323 |  | 4.05 (-1.51, 9.62), p=0.1519 |  | 0.13 (-0.50, 0.77), p=0.6849 |  |

**By sex**
| Female |  | n=275 | n=236 | n=384 | n=307 | n=9526 | n=8650 |
| --- | --- | --- | --- | --- | --- | --- | --- |
| **Pre** |  | 4.30 (0.26) | 4.47 (0.28) | 4.35 (0.23) | 4.81 (0.25) | 5.57 (5.47, 5.67) | 5.47 (5.36, 5.58) |
| **Post** |  | 2.40 (0.26) | 2.86 (0.28) | 2.48 (0.23) | 3.43 (0.25) | 3.84 (3.73, 3.94) | 3.72 (3.61, 3.82) |
| **D<sup>n</sup>** |  | -1.90 (0.28) | -1.62 (0.30) | -1.86 (0.23) | -1.38 (0.26) | -1.74 (-1.84, -1.63) | -1.75 (-1.87, -1.64) |
| **DiD (95% CI)** |  | -0.28 (-1.10, 0.53), p=0.4957 |  | -0.48 (-1.16, 0.20), p=0.1632 |  | 0.02 (-0.14, 0.17), p=0.8501 |  |
| Male |  | n=176 | n=151 | n=216 | n=184 | n=13540 | n=12495 |
| **Pre** |  | 7.74 (0.56) | 8.07 (0.60) | 7.36 (0.56) | 8.02 (0.61) | 7.99 (0.07) | 7.73 (0.07) |
| **Post** |  | 5.45 (0.56) | 4.53 (0.60) | 5.24 (0.56) | 4.67 (0.61) | 5.63 (0.07) | 5.45 (0.07) |
| **D<sup>n</sup>** |  | -2.29 (0.55) | -3.54 (0.59) | -2.12 (0.66) | -3.35 (0.72) | -2.37 (0.07) | -2.28 (0.07) |
| **DiD (95% CI)** |  | 1.25 (-0.34, 2.83), p=0.1227 |  | 1.23 (-0.69, 3.15), p=0.2097 |  | -0.09 (-0.28, 0.10), p=0.3743 |  |

Abbreviations: BP-LTCCB, brain-penetrant L-type calcium channel blockers; non-BP-LTCCB, non-brain penetrant L-type calcium channel blockers; AUD, alcohol use disorder; Pre, pre-index drinks per week; Post, post-index drinks per week; D<sup>n</sup>, change in drinks per week; DiD, difference-in-difference estimate; CI, confidence interval
Notes: average pre- and post-index date drinks per week reported as mean (standard error); light (<3 drinks per week for males and females); moderate (3-14 drinks per week for males or 3-7 drinks per week for females); heavy (>14 drinks per week for males or >7 drinks per week for females).

Subgroup analyses stratified by AUD diagnosis and sex showed no differential patterns for any contrast in either study. In the VA, there was some evidence that non-BP-LTCCB recipients and unexposed patients with heavy baseline consumption had greater decreases in drinks per week than BP-LTCCB recipients. However, this subgroup finding may reflect residual confounding and should be interpreted cautiously. In the KPNC study, sample sizes of individuals with baseline heavy drinking and AUD were small in the BP-LTCCB vs. unexposed and BP-LTCCB vs. non-BP-LTCCB comparisons. Nonetheless, we present all findings to be consistent with our preregistration.

## Discussion

This large, pre-registered study in two healthcare systems did not find evidence that BP-LTCCB initiation was associated with reduced alcohol consumption more than non-BP-LTCCB initiators or unexposed comparators to either drug class. This finding was consistent across multiple clinical and demographic subgroups, including by sex, AUD diagnosis, and baseline alcohol use. Results from this study do not support the repurposing of BP-LTCCBs for treatment of AUD or as a medication to reduce drinking behavior. Our results inform the broader literature on CCBs for addiction and other psychiatric conditions, including studies suggesting that individuals prescribed certain LTCCBs may have reduced rates of psychiatric conditions^3261^

There are multiple interpretations for these null results. It is possible that BP-LTCBBs, despite support from genetic and preclinical evidence, do not meaningfully impact brain networks associated with alcohol use and AUD. Although we analyzed participant subgroups, BP-LTCCBs may be impactful for some participant groups that were not analyzed or not assessed in this study. Notably, the KPNC sample primarily included individuals with light to moderate drinking and without AUD. Individuals prescribed BP-LTCCBs may have particular characteristics not accounted for in our study that mask potential benefits of these medications on drinking behavior.

A surprising finding was that the non-BP-LTCCB comparator, amlodipine, showed evidence of greater reductions in drinking than that reported by unexposed individuals in the VA, but not in KPNC. However, these differences were not clinically meaningful (DiDs < 0.5 across all comparisons) and may be attributable to differences unaccounted for between the two groups. Notably, a recent study that was published after the completion of the analyses presented herein, showed evidence that amlodipine has higher brain penetrability than previously thought to be the case^62^. Thus, future studies are warranted to better understand the relative permeability of the blood-brain barrier by CCBs and to parse the possible psychiatric effects of each medication. This observation suggests the brain penetrance of these drugs may not necessarily be a dichotomous yes/no definition. Nonetheless, it is important to note that, regardless of brain penetrance, this study was designed as a three-group analysis including a propensity-score matched unexposed comparator group and does not find evidence supporting that CCBs may be promising pharmacotherapeutic approaches for AUD.

To our knowledge, this is the first large EHR-based study specifically designed to analyze the impact of BP-LTCCBs on alcohol use, and the study has multiple strengths. Proposed methods and hypotheses were pre-registered, increasing transparency and limiting risks to project integrity. While prior EHR-based studies have been hindered by conduct within a single healthcare system, we conducted analyses in two different healthcare systems with complementary demographic makeup, allowing for greater diversity and generalizability. Indeed, our current findings are bolstered by having previously found converging evidence in VA and KPNC that spironolactone^63,64^ and glucagon-like peptide-1 receptor agonists^65,66^ were associated with greater reductions in alcohol use. This study is also strengthened by our ability to account for potential confounding through propensity score matching of participants in different exposure groups, and our ability to select participants whose alcohol behaviors were measured after drug prescription. Finally, unlike most large EHR datasets that rely primarily on diagnostic codes or alcohol-related events, both VA and KPNC routinely collect quantitative alcohol consumption data using validated instruments recommended by the National Institute on Alcohol Abuse and Alcoholism^50^. This allowed us to directly evaluate changes in reported alcohol use, consistent with endpoints used in randomized clinical trials.

This study has limitations. Despite our use of propensity-score matching to control for baseline differences between groups, we cannot rule out unmeasured variables that could confound the results, such as patient motivation to reduce alcohol use and participation in non-pharmacological treatments. Relatedly, LTCCBs were examined without restriction to indication. The inability to capture the clinical rationale for prescribing limited adjustment for indication-related factors, which may have introduced residual confounding by indication in contrasts with unexposed comparators. We could not consider the impact of LTCCB dose on alcohol use; if higher doses are needed to decrease alcohol use, this benefit would need to be balanced against risk for hypotension. The VA study was conducted on Veterans in care, who may not reflect the same chronic health conditions and risk behaviors as the general US population. However, these limitations are mitigated by the consistency of findings across two large healthcare systems.

In summary, this rigorous pre-registered EHR study of two health systems found that BP-LTCCBs were not associated with significantly greater reductions in drinking behavior than two comparator groups. Results from this study add further context to prior findings of potential benefit of BP-LTCCBs on psychiatric symptoms and phenotypes. Despite promising preclinical evidence, our findings suggest LTCCBs may not impact drinking behaviors at a population level. This research highlights the need for further research into other pharmacologic strategies for treating AUD.

## Data Availability

Owing to US Department of Veterans Affairs (VA) regulations and our ethics agreements, the analytic data sets used for this study are not permitted to leave the VA firewall without a data use agreement. This limitation is consistent with other studies based on VA data. However, VA data are made freely available to researchers with an approved VA study protocol. For more information, please visit https://www.virec.research.va.gov or contact the VA Information Resource Center at. The KPNC datasets of the current study are not publicly available due to potentially identifiable information (e.g., dates of diagnoses) and KPNC privacy regulations. They are available upon reasonable request, contingent on appropriate human subjects approval and data use agreements.

## Acknowledgments

This work uses data provided by patients and collected by the Department of Veterans Affairs (VA) and Kaiser Permanente Northern California (KPNC) as part of their care and support.

## Author Contributions

Study concept and design: CTR, VAP, HRK, JCG. Analysis of data: CTR, VAP. Drafting of the paper: CTR, VAP, MS, JCG. All authors provided critical revision of the paper for important intellectual content.

## Disclaimer

The contents of this publication are the sole responsibility of the authors and do not necessarily reflect the official policy or position of the Uniformed Services University of the Health Sciences, the Department of War, or Henry M. Jackson Foundation for the Advancement of Military Medicine. This research was supported in part by the Intramural Research Program of the National Institutes of Health (NIH) (MF and LL). The contributions of the NIH authors were made as part of their official duties as NIH federal employees, are in compliance with agency policy requirements, and are considered works of the United States Government. However, the findings and conclusions presented in this paper are those of the authors and do not necessarily reflect the views of the NIH or the U.S. Department of Health and Human Services.

